# Food fraud: insights from investigating a near-fatal poisoning with global implications

**DOI:** 10.1101/2023.09.27.23296221

**Authors:** Zhao Cao, Masha Yemets, Sadia Muneem, Kyle Shannon, Fatima Gowher, Jennifer Malloy, Erin Kafka, Clifford Mitchell, Joshua King, Sinisa Urban

## Abstract

Risks of food fraud have been exacerbated by ongoing supply-chain shortages and reduced regulatory oversight caused by the COVID19 pandemic. Since food fraud cases involve adulterants that are deliberately disguised, resulting poisonings are especially difficult to investigate and treat. We encountered a near-fatal poisoning with “weight-loss candlenuts” that are readily available online. We leveraged state-of-the-art high-resolution mass spectrometry (HRMS) and discovered that commonly used spectral libraries and toxin/poison standards panels do not contain the materials needed to identify the causative agents of this nearly lethal poisoning. By building new methods, we ultimately found ‘Nuez de la India’ contain high levels of uncommon cardiac glycosides, while deploying HRMS as a novel ‘chemical fingerprinting’ tool revealed them to be mislabeled yellow oleander seeds. Our work presents a rapid investigative strategy to empower future investigations, and provides guiding principles to food safety programs for treating, and indeed preventing, these potentially fatal poisonings that are increasing worldwide.

## INTRODUCTION

Food fraud results from the deliberate adulteration of foods or ingredients with superficially similar but cheaper alternatives^1,2^. The result can be an inferior product that is sold at a premium price, which cheats but otherwise does not harm consumers. Common examples include diluting high-grade honey with sugar syrups^3,4^, or cheaper fish for more desirable species^5,6^.

Cases can also result in harm when a food is adulterated with a toxic substance. Particularly widespread cases occurred when lead chromate was added to turmeric^7,8^. The result was an appealingly vibrant spice color, and, since lead chromate is heavy, a significant increase in profit (because the product is sold by weight). However, both lead and chromium are toxic metals with chronic effects and thus resulted in dangerous and sustained consumer exposures.

The COVID-19 pandemic increased the risk (albeit not necessarily the prevalence) of food fraud reaching consumers^9,10^. Increased online food ordering for direct delivery to consumers, coupled with ingredient shortages due to supply disruptions, paused or decreased facility licensure and import inspections, created a ‘perfect storm’ for criminal opportunities to exploit vulnerabilities in already stressed food safety systems^11^. The global and highly interconnected nature of modern food systems thus requires collaboration across the entire international community to address these vulnerabilities.

Food fraud becomes especially dangerous when it results in acute poisonings, because the causative agent is unknown and unsuspected, and the harm occurs rapidly. Time spent investigating the cause delays clinical action and can lead to deadly consequences. When such cases are encountered, it is especially important to mount a detailed investigation to protect public health: a specific cause must be determined, dissemination of the investigative tools and results must be made to empower testing laboratories and clinicians with the most effective course of action, and food safety regulators must be engaged with the data they require to take lawful action.

With this mission in mind, we recently conducted a complex and time-consuming investigation into the cause of a poisoning from ‘weight-loss nuts’ purchased online and sourced internationally. The resulting findings ultimately implicate food fraud as the root cause, and establish efficient methodologies for investigating such poisonings when time is strictly limited. The ultimate clinical outcome further suggests an effective course of treatment. Finally, we offer a powerful 3-dimensional ‘chemical fingerprinting’ approach to empower food safety regulators for combating food fraud more effectively, and thus avoid such poisonings in the first place.

## RESULTS

### The poisoning

An adult patient was admitted to a local hospital suffering from nausea, vomiting, and diarrhea. The patient had deliberately ingested diet nuts that were purchased online called ‘Nuez de la India’ (Fig. 1a), which is a colloquial name for candlenuts that are commonly used as a weight-loss aid. The patient’s symptoms worsened after admission, and resulted in life-threatening bradycardia (heart rate slowing) and hyperkalemia (elevated blood potassium). An electrocardiogram showed a characteristic ‘Salvador Dali mustache’ with scooping of the S-T segment.

**Fig. 1.**
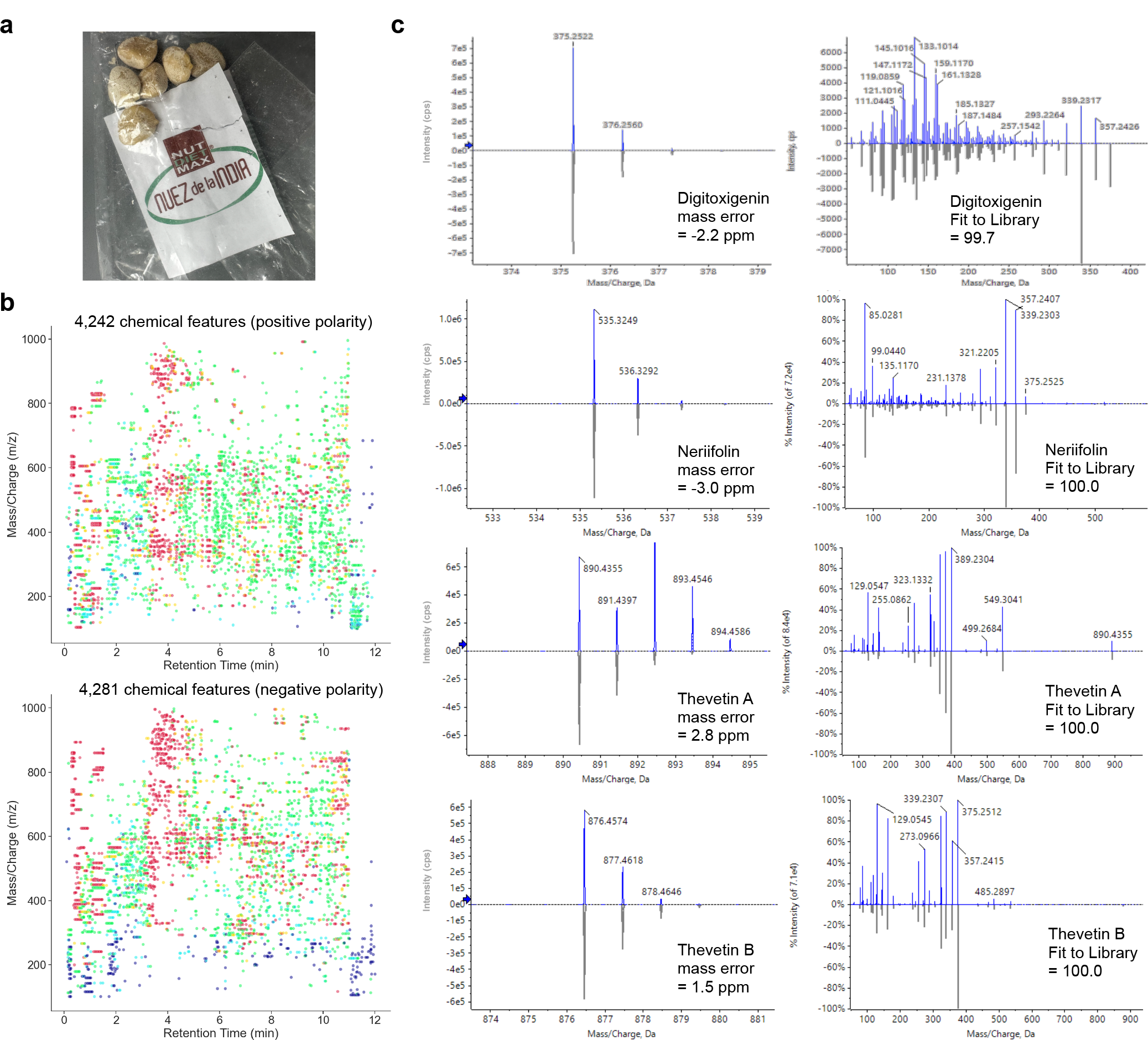
HRMS reveals Nuez de la India contain novel cardiac glycosides. **a**, Photograph of the ‘Nuez de la India’ product sample collected from the poisoned patient and delivered to the Maryland public health laboratory for investigation. **b**, All detectable ions from the Nuez de la India patient sample were graphed as accurate mass-to-charge ratio (Y-axis) versus retention time emerging from the liquid chromatography column (X-axis, in minutes), with spot color corresponding to ion abundance. **c**, Comparison of sample data (top in blue) to spectral library entries derived from known standards (mirror imaged underneath in grey). The leftmost match depicts intact accurate mass of the parent ion, plus naturally occurring isotope variant matching (smaller peaks on the right). Rightmost graphs depict experimental fragmentation pattern of intact parental ions.

The emergency physician contacted the Maryland Poison Center (MPC), and cardiac glycoside poisoning was suspected from the clinical picture^12^. Upon investigation of the case, MPC personnel discovered a recent article describing a case of suspected yellow oleander poisoning^13,14^ and promptly administered anti-digoxin antidote^15^. This intervention resulted in rapid improvement in bradycardia. By 12 hours post-admission the patient’s heart rate normalized, and the next day the patient was released from the hospital in a normal state of health.

### Current methods have dangerous blind spots

The suspected weight-loss nuts were collected from the patient and submitted to the Maryland public health laboratory to determine the cause of the poisoning. Since fundamentally this was an unknown, we deployed high-resolution mass spectrometry (HRMS) that has the power to resolve and characterize all chemicals present in a complex food sample in a low-bias manner^16^.

HRMS investigations typically involve two phases. First, all detectable chemicals in a sample are resolved and their mass is measured with high accuracy (usually to 4-decimal places). Next, the resulting ‘chemical features’ of intact and experimentally fragmented chemicals are then compared to those in a reference spectral library built from known standards in order to identify the individual chemicals present in the sample.

Scans of the Nuez de la India in both the positive and negative polarity resolved as many as ∼8,500 chemical features (Fig. 1b). We used this dataset to interrogate a large National Institute of Standards and Technology (NIST) library of >17,500 compounds, but only found evidence of a single poison, namely digitoxigenin (Fig. 1c). However, since the poisoning was suspected to be from cardiac glycosides, we subsequently purchased several additional standards, recorded their spectra, and added them to our library^17,18^. This proved to be a key point.

Enhanced spectral library searching revealed some of the most abundant chemicals in the Nuez de la India to be digitoxigenin-3-α-L-thevetoside (neriifolin), thevetin A, and thevetin B (Fig. 1c). Importantly, further analysis suggested that the digitoxigenin we initially detected formed as a breakdown product of neriifolin^19^. As such, none of the cardiac glycosides that were responsible for the poisoning were in the widely used NIST spectral library.

As a Chemical Emergency Preparedness & Response Laboratory, we also had at our disposal a targeted (non-HRMS) high-sensitivity mass spectrometry method that utilizes 44 poison/toxin standards to detect and quantify common poisons/toxins found in foods. This orthogonal approached further revealed that other more common cardiac glycosides including digoxin, digoxigenin, and digitoxin were not present in the Nuez de la India, but the three cardiac glycosides that were responsible for the poisoning also were not identified because none were present in this ‘go-to’ food toxins/poisons method.

Finally, we performed targeted analysis with chemical standards to confirm and quantify neriifolin, thevetin A, and thevetin B that we identified using HRMS. This approach revealed these cardiac glycosides to be present at a sum of >20 mg/g in the Nuez de la India product (Fig. 2). This extraordinarily high level likely explains the rapid and dramatic poisoning the patient experienced.

**Fig. 2.**
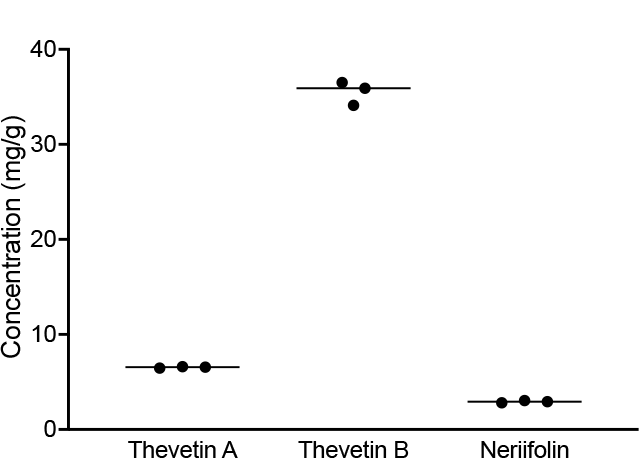
Nuez de la India contain extraordinarily high levels of neriifolin, thevetin A, and thevetin B. Quantification of cardiac glycosides in multiple reaction mode analyzed in triplicate with known standards. Horizontal line depicts the mean of each dataset.

### Poisoning reoccurrence is likely

The core mission of a public health laboratory is to protect public health prospectfully. We therefore next sought to determine whether this type of poisoning could happen again by purchasing and testing a new package of the product from the same online retailer as the case patient.

The first question we asked is whether the patient sample is chemically different from the new product, and leveraged the power of HRMS to record large datasets of distinct chemicals in three dimensions (accurate mass-to-charge, column retention time, and ion abundance) even when the detected chemicals are not identified. HRMS revealed that the new product had a ‘chemical fingerprint’ that is indistinguishable from the nuts ingested by the patient across all >6,300 chemical features detected in both negative and positive polarity analyzed in parallel (Fig. 3).

**Fig. 3.**
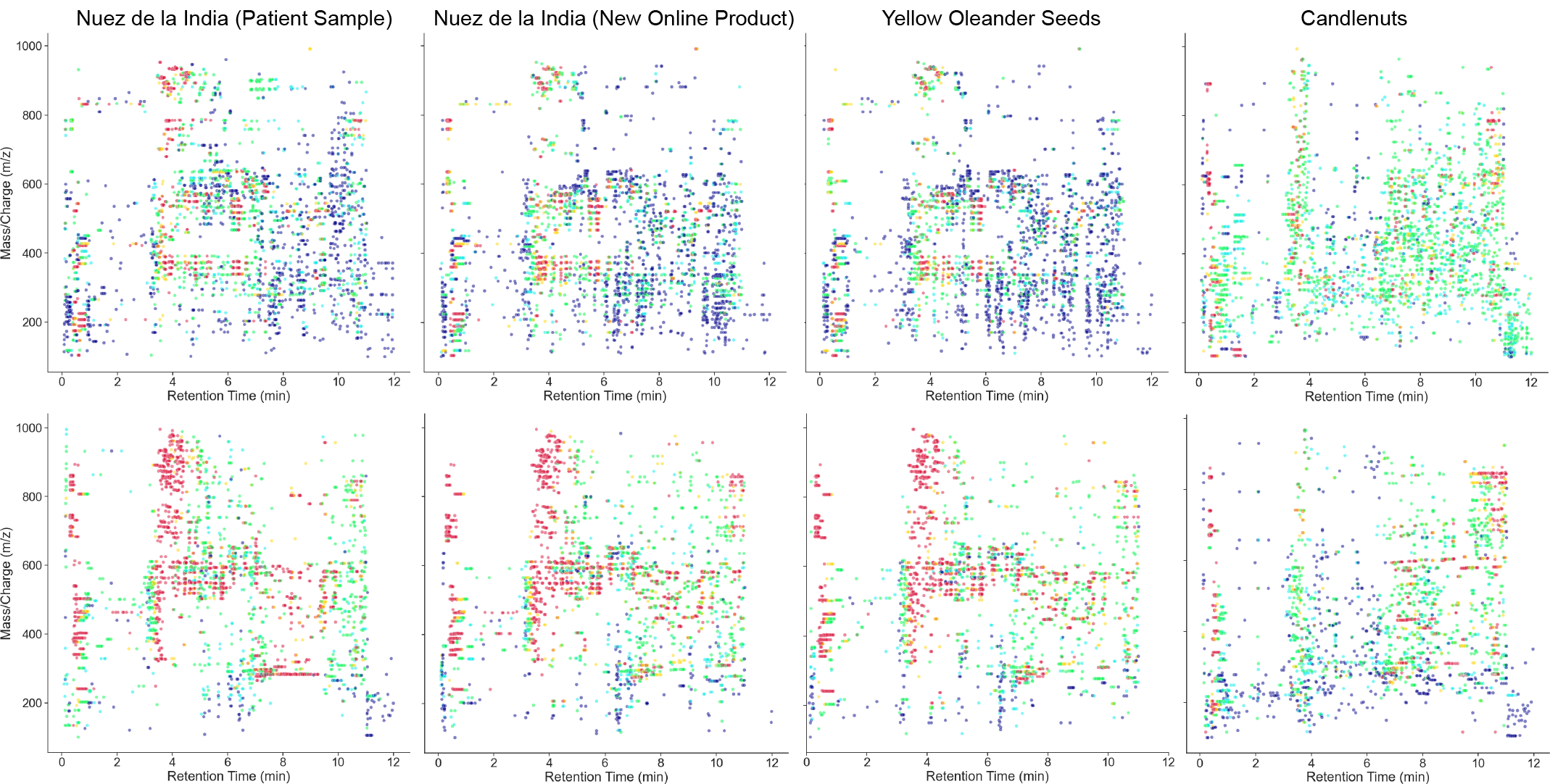
HRMS reveals Nuez de la India are mislabeled yellow oleander seeds. Distribution of all detectable ions (accurate mass-to-charge ratio on Y-axis versus retention time on X-axis, with spot color corresponding to ion abundance) in both positive (top) and negative (bottom) modes serve as ‘chemical fingerprints’ for each nut type.

We then turned to our targeted method to confirm and quantify the three key cardiac glycosides we ultimately discovered in the patient samples that likely led to the poisoning. The new product also proved to have extraordinarily high levels of the same cardiac glycosides as the patient sample (Fig. 4). Thus, readily available product not only could, but in fact will, exert the same harm to other unsuspecting dieters.

**Fig. 4.**
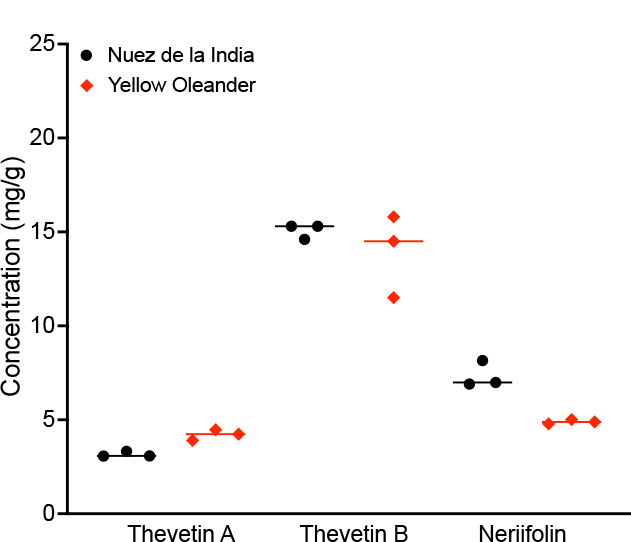
Principle component analysis confirms Nuez de la India are mislabeled yellow oleander seeds. Principal component analysis of all nut samples revealed strong clustering of Nuez de la India from both the patient and online retailer with bona fide yellow oleander seeds analyzed in triplicate, but away from candlenuts and other nut types. Graphed are the largest three eigenvectors emerging from the analysis of these 22 HRMS datasets.

### Food fraud is the true cause

The final question we asked was why this suite of cardiac glycosides is present in these purported Nuez de la India weight-loss candlenuts in the first place. We thus purchased known candlenuts and yellow oleander seeds (which were suspected to be the cause of the poisoning) from horticulturalists and again compared their ‘chemical fingerprints’ by running HRMS in both polarities. Remarkably, the ‘chemical fingerprint’ of the Nuez de la India resulting from >6,400 chemical features in 3-dimensions was indistinguishable from that of yellow oleander seeds, but obviously different from that of bona fide candlenuts (Fig. 3).

To apply an additional statistical evaluation to these complex, 3-dimensional datasets, we compared HRMS spectra run in triplicate using principal component analysis, which independently revealed that the Nuez de la India cluster tightly with yellow oleander seeds, and far away from candlenuts or any of the other 6 different nut controls that we analyzed (Fig. 5). Moreover, yellow oleander seeds contained neriifolin, thevetin A, and thevetin B as expected^17,18^, and at levels indistinguishable from the Nuez de la India that we analyzed in parallel (Figure 4).

**Fig. 5.**
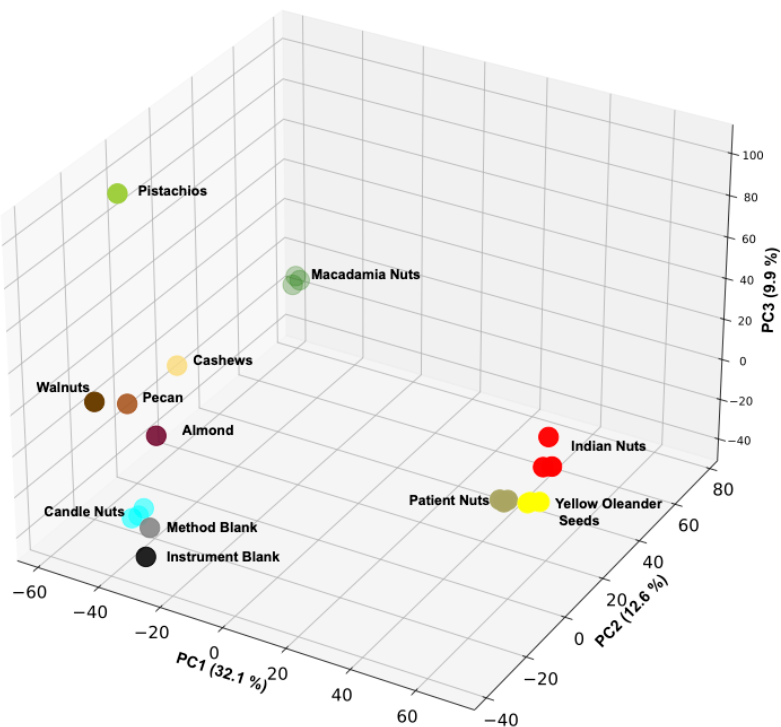
Nuez de la India contain levels of cardiac glycosides indistinguishable from yellow oleander seeds. Comparison of neriifolin, thevetin A, and thevetin B levels in Nuez de la India and yellow oleander seeds determined in triplicate in multiple reaction mode with known standards. Horizontal line depicts the mean.

Importantly, the abundance of all detected chemicals were indistinguishable between pure yellow oleander seeds and the Nuez de la India (Fig. 3, 4, 5), revealing that Nuez de la India are not just contaminated with some yellow oleander seeds, but are themselves entirely yellow oleander seeds. We therefore conclude that the Nuez de la India are, in fact, the result of food fraud: the product is entirely comprised of yellow oleander seeds that are mislabeled, and not at all candlenuts as the name Nuez de la India indicates.

## DISCUSSION

Food fraud is especially sinister when foods are adulterated with a disguised ingredient that is acutely poisonous. The most effective way for food systems to address these cases is to mount a thorough investigation, build tools that empower future responses, adapt regulatory systems to accept new analytical tools, and disseminate the results widely across disciplines.

We leveraged HRMS to investigate a local cardiac poisoning event that resulted from food fraud from a product sourced internationally. In the process of this work, we discovered that the commonly used spectral libraries and poison/toxin standards are unable to identify the poisons responsible. Nevertheless, applying HRMS as a research tool ultimately proved to be successful in identifying cardiac glycosides in highly complex samples, and to discern the true identity of the mislabeled yellow oleander seeds using HRMS datasets as ‘chemical fingerprints’.

While we were ultimately successful in this investigation, the first key take-away to note is that current tools at the disposal of most laboratories are unlikely to be effective when investigative speed is paramount. As such, a key goal of this article is to disseminate our applied research in order to help equip laboratories and clinicians for investigating future poisonings. We thus urge laboratory readiness programs including FDA’s Food Emergency Response Network (FERN) and CDC’s Laboratory Response Network – Chemistry (LRN-C) to address this debilitating gap by adding these agents to the methods and libraries that they maintain and transfer to state and local laboratories.

It should not be underestimated just how deadly the consequences of spending time to gather the required materials and build the appropriate mass spectrometry methods could be: while the clinical outcome of our case patient had a happy ending (before we could perform any chemical analyses, which took several months to complete), a prior case judiciously reported in 2020 resulted in a fatality. A patient in Minnesota ingested just 5 Nuez de la India, was admitted to a local hospital, and suffered a cardiac arrest^13^. She was resuscitated and placed in an intensive care unit, but died 2 hours later while her clinical team was diligently working on a treatment.

Our experience also offers guidance to clinicians treating Nuez de la India poisonings. Our patient had ingested 12 nuts, and exhibited hyperkalemia of 5.9 (untreated levels >5.5 are generally lethal in cardiac glycoside poisonings^13-15^), but was administered 8 vials of digoxin antibody fragments promptly and survived. This is, to our knowledge, the first documented case of a successful clinical treatment for a Nuez de la India poisoning event, and thus provides a precedent for future life-saving responses: even a high dose of 12 nuts can be countered with as few as 8 anti-digoxin vials, if administered quickly. Nevertheless, even with this successful course established, two main challenges remain.

First, the antidote is expensive (listed online as $4,666 USD per vial), and generally not stocked by hospitals in large quantities. Our local hospital had 8 vials on hand, and administered all in order to treat the patient. The tragic case in Minnesota is especially sobering: while the patient was in the intensive care unit, her clinical team found their facility had only 5 vials on hand and was calling other hospitals to secure additional doses (because treating yellow oleander poisoning is thought to require 10-20 times the regular dose^15^) when the patient suddenly died^13^. In the end, it was discovered that the entire city had no more than 20 doses on hand, which may be enough to save only one or two poisoned residents.

The final and perhaps most difficult challenge is concerns food safety systems: the Nuez de la India product is still readily available internationally despite multiple poisonings and with no warning of a possible fatality. In fact, the product insert recommends dosage of one-quarter of one nut, but is written nearly entirely in Spanish, and the nuts are small and likely to be consumed whole and successively. Swift and decisive regulatory action is thus required internationally, but the difficulty in connecting even robust investigative tools such as chemical fingerprinting to existing legal frameworks remains a serious roadblock.

In fact, it is surprising just how vastly underused chemical fingerprinting is in food fraud case investigations^16,20^: while HRMS is exquisitely suited to address the challenges of food fraud, food safety programs, including our local regulatory programs, rarely accept its findings. Part of this discrepancy stems from food safety statutes that were written when this powerful technology was experimental or not readily available. As such, satisfying outdated statutes that lead to regulatory action with 3-dimensional HRMS datasets requires ‘fitting a square peg into a round hole’, and renders the powerful datasets moot. Food safety regulators should therefore work to adapt well meaning but outdated statutes to modern analytical approaches that have the power to solve dangerous cases of food fraud quickly, thereby allowing the data to be used seamlessly within a robust legal framework that can stop food fraud at its source.

In summary, it is our hope that dissemination of this food fraud case in all of its complex chemical, clinical, and regulatory dimensions helps clinicians and laboratorians take the best course of action to investigate and treat these potentially fatal poisonings, alerts food safety programs to the intrinsic risk of this product that can easily be fatal, and enables regulators to take swift and decisive action to prevent these deadly poisonings that are already becoming more common across the globe.

## ONLINE METHODS

### Samples and Standards

All nuts were received at the Maryland Department of Health Laboratories Administration under strict Chain of Custody protocols, and kept locked in a secure location at room temperature until use. Nut Diet Max brand ‘Nuez de la India’ was purchased through Amazon, while pistachio, macademia, walnut, pecan, cashew, and almond nut controls were collected from retail, by Maryland’s Office of Food Protection. From horticulturalists were purchased yellow oleander seeds (4 seeds of *Thevetia peruviana*, item # 334337769995, The Plant Attraction) and candlenuts (25 unpolished seeds of *Aleurites moluccanus*, BabuBotanicals), which were then dissected in the laboratory to release the seeds from the nuts for extraction.

Standards were purchased from the following sources: neriifolin (Sigma Cat. No. SMB01017), thevetin A (Sigma Cat. No. SMB00166), thevetin B (Cat. No. CFN96147 from AOBIOUS), digoxigenin (Sigma Cat. No. D9026-100mg), digitoxigenin (Sigma Cat. No. D9404-100mg), digitoxin (Cerilliant Cat. No. D-067), digoxin (Cerilliant Cat. No. D-029), and custom poisons/toxins standards mix designed for laboratories in FDA’s Food Emergency Response Network (o2si Cat. No. G34-140341-KIT).

### Sample Extraction

Each nut type was homogenized in a Ika A10 Basic blender, and 2 g of the resulting homogenate was transferred to a 50mL disposable centrifuge tube. 2 ml of MilliQ water was added to the samples, vortexed, and allowed to soak for at least 2 min at room temperature. Internal standards (250 μl of 20 μg/ml tri-phenyl phosphate for positive mode, and 100 μl of 3 ppm ^13^C_4_-perfluorooctane sulfonate for negative mode) were added to each sample, blank, and matrix spikes to allow internal monitoring of recovery and signal normalization.

Each extraction batch included a method blank in which nuts were excluded, and a matrix spike in which nuts were spiked to 500 ppb of each target analyte to assess recovery. Dietary supplements were included as the initial extractions as negative controls, while macademia nuts and candlenuts served this purpose for later batches.

Following the aqueous soak and internal standards addition, 9.65 ml of acetonitrile was added to the samples and matrix blank, 9.4 mL to the matrix spike, and 10 mL to the reagent blank, and shaken for at least 1 minute prior to centrifugation at 2,800 g for 10 min in a Thermo Scientific Legend XT centrifuge equipped with a swinging-bucket rotor (75003180/75003608). Supernatants were filtered through a 0.2 μm nylon filter (Pall Acrodisc AP-4436) into a fresh tube, and aliquots were transferred to autosampler vials for analysis by mass spectrometry.

### High-Resolution Mass Spectrometry

Nut extracts were run on an AB Sciex QTOF X500R high-resolution mass spectrometer equipped with a AB Sciex/Shimadzu EXION-LC 2.0 ultra-high pressure liquid chromatograph (UPLC). 2 μl of each extract was injected onto a C18 column (Waters Acquity UPLC BEH 1.7 μm, 50 × 2.1 mm) and run with the following resolving conditions:

| Time (min) | 2nM NH<sub>4</sub>OAc in 95:5 water:acetonitrile (%) | Acetonitrile (%) | Flow rate (ml/min) |
| --- | --- | --- | --- |
| 0.0 | 98 | 2 | 0.35 |
| 0.2 | 98 | 2 | 0.35 |
| 4.0 | 70 | 30 | 0.40 |
| 7.0 | 45 | 55 | 0.40 |
| 9.0 | 25 | 75 | 0.40 |
| 10 | 5 | 95 | 0.40 |
| 10.4 | 98 | 2 | 0.40 |
| 12.0 | 98 | 2 | 0.35 |

High-resolution data was collected using Information-Dependent Acquisition (IDA) with the following parameters:

| Parameter | Positive polarity | Negative polarity |
| --- | --- | --- |
| Method duration | 12 min | 12 min |
| Total scan time | 1.574 s | 1.574 s |
| Estimated cycles | 457 | 457 |
| Ion source gas 1 / 2 | 50 psi | 50 psi |
| Curtain gas | 30 psi | 30 psi |
| CAD gas | 7 psi | 7 psi |
| temperature | 500 °C | 500 °C |
| Spray voltage | 5500 V | -4500 V |
| TOF-MS range (m/z) | 100-1000 | 100-1000 |
| TOF-MS accumulation time | 0.1 s | 0.1 s |
| Declustering potential | 50 V | -80 V |
| DP spread | 0 V | 0 V |
| Collision energy | 10 V | -10 V |
| CE spread | 0 V | 0 V |
| TOF-MSMS range (m/z) | 50-1000 | 50-1000 |
| TOF-MSMS accumulation time | 0.1 s | 0.1 s |
| Declustering potential | 50 V | -80 V |
| DP spread | 0 V | 0 V |
| Collision energy | 35 V | -35 V |
| CE spread | 0 V | 0 V |
| Maximum candidate ions | 14 | 14 |
| Intensity threshold | 10 cps | 10 cps |
| Dynamic background subtraction | Active | Active |

The resulting high-resolution mass spectral datasets were processed in AB Sciex OS v3.0.0.3339 Acq Qual/Quant software (Cat. No. 5301606) using a AB Sciex All-in-One 2.1 NIST 2017 library (Cat. No. 5084472) for the identification of unknowns, to which we manually added experimental spectra for neriifolin, thevetin A, and thevetin B. Data was plotted in AB Sciex OS software (for library matching data) and Python Jupyter Notebook (for displaying all datasets in Figure 1a and 2a, with the following color scheme: 0-499=Blue, 500-999=Turquoise, 1000-4999= Green, 5000-9999=Yellow, 10,000+=Red.

### Targeted Poison/Toxin Screening and Quantitation

Nut extracts were run on a high-sensitivity Sciex QTRAP 6500+ tandem mass spectrometer equipped with an Agilent Infinity 1290 high pressure liquid chromatograph (HPLC). 1 μl of each extract was injected onto a Zorbax SB C-18, 2.0 × 150 mm, 1.8 μm, column (Agilent Cal. No. 883700-922) and run with the following resolving conditions:

| Time (min) | 0.1% (v/v) formic acid in water | 0.1% (v/v) formic acid in acetonitrile | Flow rate (ml/min) |
| --- | --- | --- | --- |
| 0 | 95 | 5 | 0.2 |
| 15 | 65 | 35 | 0.2 |
| 20 | 5 | 95 | 0.2 |
| 20.1 | 5 | 95 | 0.3 |
| 25.0 | 95 | 5 | 0.3 |
| 25.1 | 95 | 5 | 0.2 |
| 32.0 | 95 | 5 | 0.2 |

Mass spectrometry was run in positive polarity multiple-reaction mode (MRM) with the following ion transitions and parameters:

| Analyte | Q1 (m/z) | Q3 (m/z) | RT (min) | DP (Volts) | CE (Volts) |
| --- | --- | --- | --- | --- | --- |
| thevetin A | 874.3 | 550.2 | 12.8 | 19 | 35 |
|  | 890.1 | 389.1 | 12.8 | 36 | 17 |
| thevetin B | 876.5 | 375.2 | 14.7 | 1.0 | 53 |
|  | 876.5 | 339.2 | 14.7 | 1.0 | 31 |
| neriifolin | 535.3 | 339.2 | 18.7 | 126 | 21 |
|  | 535.3 | 375.2 | 18.7 | 126 | 17 |

For quantification, instrument performance was calibrated using 7-point calibration standards ranging in concentration from 0.1 ppm to 20 ppm, resulting in linear calibration curves not forced through zero, using 1/x weighting, and R^2^ values exceeding 0.99. Extracts were then run in triplicate to quantify target analytes. Owing to the extraordinarily high levels of cardiac glycosides in Nuez de la India and yellow oleander seeds, extracts had to be diluted 1,000-fold to be within the quantifiable range. Data was graphed in GraphPad Prism 9.4.0.

### Principal Component Analysis

Positive-mode high-resolution mass spectrometry datasets were analyzed for each nut type with respective method and instrument controls by principal component analysis in the MarkerView v1.3 software suite (AB Sciex Cat. No. 5044846). Nuez de la India, candlenuts, and macademia nuts were extracted and run in triplicate (biological replicates), while owing to sample limitations yellow oleander seeds and the patient Nuez de la India were analyzed in triplicate (technical replicates). PCA was performed with logarithmic weighting and pareto scaling, resulting in eigenvalues of the three major components (eigenvectors) accounting for 32.1% (PC1), 12.6% (PC2), and 9.9% (PC3), which were then graphed in 3-dimensions using Python Jupyter Notebook.

## Data Availability

All data produced in the present work are contained in the manuscript.

## Author contributions

Z.C. conducted all analytical chemistry experiments and data analysis, S.U. designed and oversaw the study, S.M. contributed to data interpretation, F.G. graphed the data, K.S. and E.K. collected all samples, M.Y., J.M., J.K., and C.M. analyzed the patient poisoning information, S.U. wrote the manuscript and made the figures, all authors commented on and approved the final submission.

## Data sharing

All data required to support the conclusions of this work are included directly within the article. Relevant raw data are available from the corresponding author upon request. Please note that all such requests need to provide a sound methodological justification, and will require approval from the State of Maryland.

## Acknowledgements

We are grateful to CDC for their support through the LRN-C network, FDA for helpful discussions and their support through the FERN network, EPA for a ROAR collaboration on HRMS method development for non-targeted analysis, and AB Sciex for technical and method support. We also extend our sincere appreciation to the Association of Public Health Laboratories for funding that supported F.G. as an APHL/CDC Informatics Fellow, and for competitively funding our high-resolution mass spectrometer purchase.

